# Diagnostic Value of Large Language Model-Extracted Gross Brain Findings in Neurodegenerative Diseases

**DOI:** 10.64898/2026.09.12.26362866

**Authors:** Daisuke Ono, Dennis W. Dickson, Shunsuke Koga

**Affiliations:** Department of Neurology and Neurological Science, Institute of Science Tokyo, Tokyo, Japan; Department of Neuroscience, Mayo Clinic, Jacksonville, Florida, USA; School of Statistical Thinking, The Institute of Statistical Mathematics, Tokyo, Japan; Department of Pathology and Laboratory Medicine, Hospital of the University of Pennsylvania, Philadelphia, Pennsylvania, USA

**Keywords:** machine learning, large language model, brain bank, autopsy, neurodegenerative disease, neuropathology

## Abstract

Gross brain findings help guide differential diagnosis, but their diagnostic value across major neurodegenerative diseases remains incompletely characterized. We evaluated whether gross descriptions in autopsy reports could classify cases by final neuropathologic diagnosis. We analyzed 5,613 autopsy cases from the Mayo Clinic Brain Bank collected between 1998 and 2023, including seven diagnostic categories: Alzheimer disease (AD), Lewy body disease (LBD), combined AD and LBD (AD-LBD), frontotemporal lobar degeneration, corticobasal degeneration, progressive supranuclear palsy (PSP), and multiple system atrophy (MSA). A fine-tuned large language model (LLM) converted narrative gross descriptions into semi-quantitative scores for 39 features. Extraction accuracy was 0.95 in 200 manually annotated feature-level test examples. A CatBoost classifier used the extracted scores as input, whereas a second fine-tuned LLM used standardized textual descriptions of the same features. Both classifiers also included age at death, sex, and brain weight. Performance was evaluated in the same held-out test set of 562 cases. CatBoost achieved an accuracy of 0.73, a Cohen’s kappa of 0.65, and a macro-average area under the receiver operating characteristic curve of 0.92. The text-based LLM achieved an accuracy of 0.75 and a kappa of 0.68. Macro-average sensitivity was 0.66 for both models. PSP sensitivity was 0.92 with CatBoost and 0.93 with text-based LLM. Corresponding sensitivities were 0.87 and 0.92 for MSA, but only 0.21 and 0.03 for AD-LBD. Feature attribution analysis identified subthalamic nucleus atrophy and putaminal abnormalities as contributors to PSP and MSA predictions, respectively. Narrative gross descriptions contained diagnostic information, particularly for disorders with distinctive macroscopic patterns, whereas combined AD-LBD remained difficult to distinguish.

## Introduction

Neuropathologic assessment at autopsy remains essential for accurate diagnosis of neurodegenerative diseases. Histopathology and immunohistochemistry establish the definitive diagnosis and are used to evaluate the accuracy of clinical criteria, imaging findings, and fluid biomarkers.[6, 26] In routine autopsy practice, clinical information and antemortem imaging provide essential context, but gross examination of the brain remains a critical step in refining the differential diagnosis before microscopic slides are reviewed. Brain weight, regional cortical and subcortical atrophy, ventricular enlargement, and pigment loss in the substantia nigra and locus coeruleus are evaluated during gross examination and can narrow the differential diagnosis.[1, 9] These macroscopic observations are also linked to antemortem imaging because structural neuroimaging often reflects tissue loss and regional vulnerability that later become visible at autopsy.[21]

Several neurodegenerative diseases have recognizable gross patterns. Progressive supranuclear palsy (PSP) often shows midbrain and subthalamic nucleus atrophy, superior cerebellar peduncle involvement, dentate nucleus changes, and substantia nigra pigment loss.[5] Multiple system atrophy (MSA) is associated with putaminal atrophy or discoloration, pontine atrophy, cerebellar atrophy, and cerebellar white matter degeneration.[35] Corticobasal degeneration (CBD) may show asymmetric frontoparietal cortical atrophy with variable basal ganglia and nigral involvement,[4] whereas Alzheimer disease (AD), frontotemporal lobar degeneration (FTLD),[23] and Lewy body disease (LBD) differ in the prominence and distribution of cortical and medial temporal atrophy.[1, 15, 25] Although these patterns are widely taught in neuropathology, their diagnostic value has rarely been quantified systematically across large autopsy cohorts.

Large language models (LLMs) provide a practical approach to converting narrative clinicopathologic text into structured variables, an important need in brain bank research where gross and microscopic findings are often stored as non-standardized report text. Stroganov et al. showed that LLMs can extract structured macroscopic and microscopic observations from unstructured neuropathological reports in the NIH NeuroBioBank.[33] Recent Mayo Clinic Brain Bank studies have also used fine-tuned GPT models to structure longitudinal clinical records for data-driven clinicopathologic analyses, including early PSP subtyping and prediction of neuropathologic diagnoses in parkinsonism using CatBoost-based models.[28, 29]

The present study extends these approaches to autopsy gross descriptions. We used an LLM to convert narrative gross descriptions into semi-quantitative scores for 39 macroscopic features and tested whether these variables could predict seven neuropathologic diagnoses using a table-based CatBoost classifier and a text-based LLM. Our goal was to characterize disease-associated macroscopic patterns and assess diagnostic classification using these report-derived features.

## Materials and Methods

### Case selection

This retrospective study used autopsy cases from the Mayo Clinic Brain Bank for Neurodegenerative Disorders. Autopsies were performed after consent from the legal next of kin or an individual with legal authority to grant permission for autopsy. We identified 7,379 cases with available gross brain descriptions collected between 1998 and 2023. For the purpose of this study, which aimed to evaluate the impact of macroscopic findings on the diagnosis of major neurodegenerative diseases, we excluded infrequent diagnostic categories comprising fewer than 3% of the total cases, such as amyotrophic lateral sclerosis (ALS) (n = 219), major cerebrovascular disease (n = 149), and coexisting PSP and AD (n = 83). Pick disease (n = 44) was also excluded as an infrequent diagnostic category. After excluding 1,766 cases, 5,613 cases were included in the final analysis (**Fig. 1**).

**Fig. 1.**
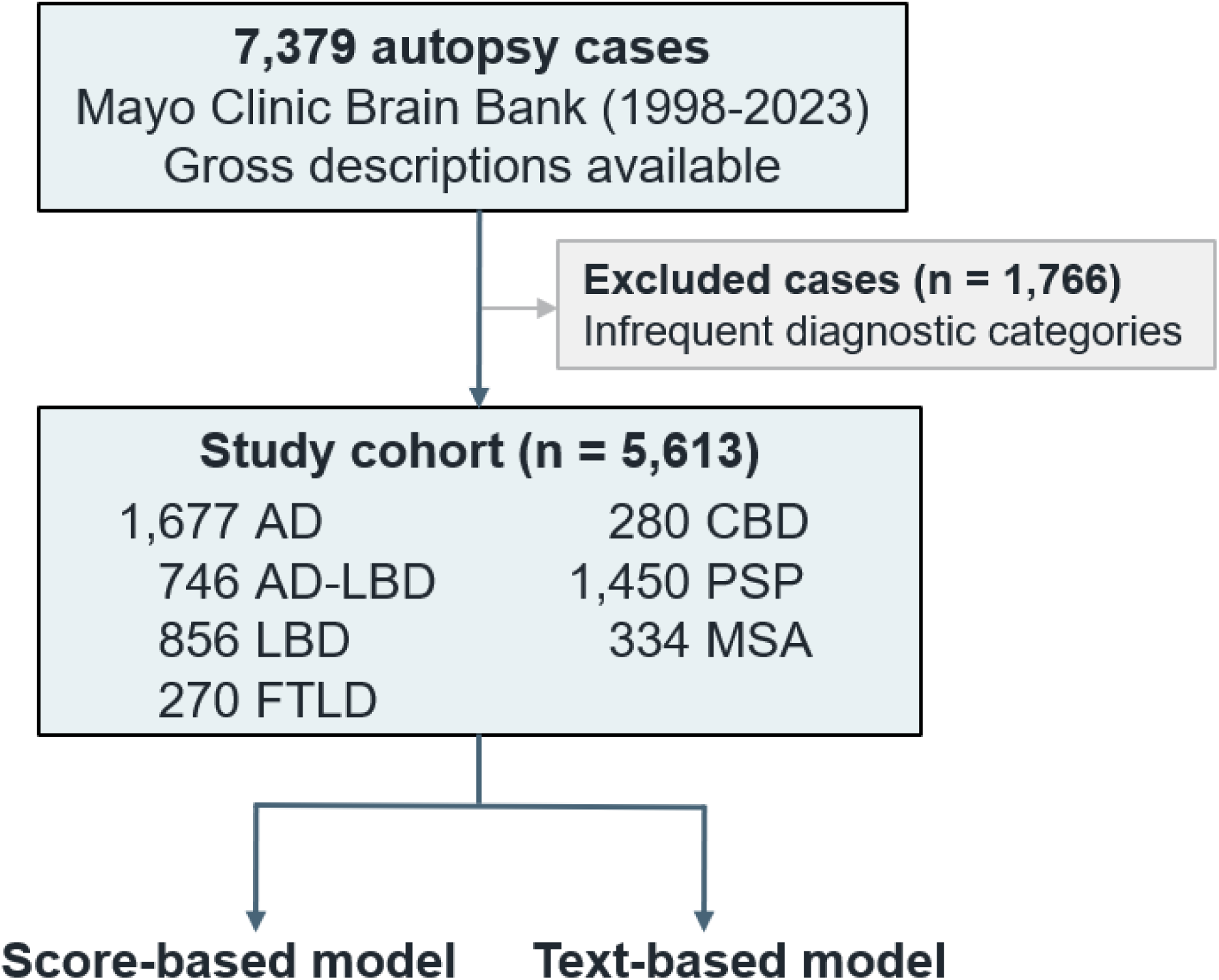
Study cohort selection and model development. A total of 7,379 Mayo Clinic Brain Bank autopsy cases with gross findings from 1998 to 2023 were identified. After excluding 1,766 cases with infrequent diagnostic categories, 5,613 autopsy-confirmed cases representing seven major neuropathologic diagnoses were included for the development of score-based and text-based classification models. Abbreviations: AD, Alzheimer disease; AD-LBD, Alzheimer disease with Lewy body disease; CBD, corticobasal degeneration; FTLD, frontotemporal lobar degeneration; LBD, Lewy body disease; MSA, multiple system atrophy; PSP, progressive supranuclear palsy.

This retrospective study used autopsy cases from the Mayo Clinic Brain Bank for Neurodegenerative Disorders. Consent for autopsy was obtained from the legal next of kin or another legally authorized representative. The Institutional Review Board of Mayo Clinic gave ethical approval for this work (IRB 24-006694). The board waived the requirement for additional informed consent for this retrospective study.

### Demographic, clinical, and genetic data

Demographic and clinical information, including age at death, sex, disease duration, and antemortem clinical diagnoses, was extracted from the Mayo Clinic brain bank database. Disease duration was defined as the interval from symptom onset to death. Multiple clinical diagnoses were retained when recorded. Previously determined APOE genotypes and MAPT H1/H2 haplotypes were retrieved from the brain bank database when available. APOE ε4 and MAPT H2 carrier status indicated the presence of at least one ε4 allele or H2 haplotype, respectively. The genotyping methods used in this brain bank have been described previously.[16]

### Neuropathologic assessment

Under the brain bank protocol, one cerebral hemisphere was frozen and the contralateral hemisphere was fixed in formalin for neuropathologic assessment. Formalin-fixed tissue underwent standardized sampling and neuropathologic evaluation as previously described in Mayo Clinic Brain Bank studies.[16, 32] Regions typically sampled included multiple neocortical regions, hippocampus, amygdala, basal ganglia, thalamus, subthalamic nucleus, midbrain, pons, medulla, and cerebellum including the dentate nucleus. Routine hematoxylin and eosin staining and thioflavin S fluorescence microscopy were performed in all cases. Additional immunohistochemistry for phosphorylated tau, alpha-synuclein, beta-amyloid, phosphorylated TDP-43, or fused in sarcoma (FUS) was performed as needed.[32]

All diagnoses were rendered by an experienced neuropathologist (D.W.D.). Braak neurofibrillary tangle stage and Thal amyloid phase were assigned using established criteria, and AD neuropathologic change was assessed according to NIA-AA criteria.[26] AD was defined using previously reported operational criteria as Braak stage ≥III and Thal phase ≥3.[32, 36] LBD was evaluated using alpha-synuclein immunohistochemistry and established consensus approaches.[25] In the present study, diffuse, transitional, and brainstem types were categorized as LBD, whereas incidental and amygdala-predominant Lewy bodies were not. Incidental Lewy body pathology denotes Lewy bodies without significant associated neuronal loss or clinical manifestations attributable to LBD.[17] AD-LBD denoted coexisting AD and LBD under these operational criteria. PSP and CBD were diagnosed based on distribution and morphology of 4-repeat tau pathology, including tufted astrocytes in PSP and astrocytic plaques in CBD.[5, 15, 16] MSA was diagnosed based on neuronal loss and gliosis in striatonigral and olivopontocerebellar systems with alpha-synuclein-positive glial cytoplasmic inclusions.[35] The FTLD group comprised 241 cases of FTLD-TDP, 21 cases of FTLD with FUS pathology, five cases of unclassifiable FTLD-tau, and three cases of unclassifiable FTLD. FTLD-TDP and FTLD-FUS were diagnosed based on characteristic patterns of neurodegeneration and TDP-43- or FUS-immunoreactive pathology, respectively.[3, 23] The five FTLD-tau cases did not meet neuropathologic criteria for PSP or CBD and were not classified as MAPT mutation-associated FTLD. The three unclassifiable FTLD cases lacked tau-, TDP-43-, and FUS-immunoreactive pathology.[3, 27] Data on cerebrovascular disease (CVD) and limbic TDP-43 pathology were retrieved from the brain bank database. CVD assessment included infarcts, hemorrhages, ischemic tissue changes, arteriolosclerosis, atherosclerosis, and cerebral amyloid angiopathy, evaluated by routine histology and thioflavin S staining.[19] Limbic TDP-43 pathology was assessed by immunohistochemical screening of the amygdala and recorded as present or absent.[37]

### Scoring of macroscopic findings using an LLM

Model inputs comprised age at death, sex, brain weight, and 39 gross brain findings extracted from autopsy reports. The gross findings included cortical atrophy, frontal lobe atrophy, parietal lobe atrophy, temporal lobe atrophy, occipital lobe atrophy, hippocampal formation atrophy, amygdala atrophy, ventricular enlargement, caudate atrophy, putamen atrophy, putamen discoloration, globus pallidus atrophy, subthalamic nucleus atrophy, midbrain atrophy, pontine atrophy, superior cerebellar peduncle atrophy, dentate nucleus atrophy, cerebellar white matter atrophy, substantia nigra pigment loss, locus coeruleus pigment loss, and related macroscopic variables (**Supplementary Table S1**). These findings were originally recorded in narrative form in the gross description section of the neuropathology report. The input text contained no diagnostic labels or microscopic findings.

A fine-tuned LLM was used to convert narrative gross descriptions into semi-quantitative numerical variables. A neuropathologist (S.K.) manually scored 2,000 feature-level examples from gross descriptions across the 39 macroscopic features using feature-specific scales ranging from 0 to 1, 0 to 2, or 0 to 3 (Supplementary Table S1). The annotated excerpts were divided into 1,800 for fine-tuning and 200 for testing. The GPT-4o-mini model (gpt-4o-mini-2024-07-18) was fine-tuned for three epochs with a batch size of 1 and a learning rate multiplier of 2, using the prompt provided in Supplementary Table S2. In the extraction test set of 200 feature-level examples, the fine-tuned LLM achieved an extraction accuracy of 0.95 and a Cohen’s kappa of 0.92, demonstrating high agreement with the neuropathologist’s ratings. Based on this performance, the fine-tuned model was applied to the study dataset to assign a score to each macroscopic feature. NA outputs were treated as absent under the assumption that gross abnormalities would have been documented during systematic examination. The resulting tabular dataset was used for score-based classification.

### Score-based classification using CatBoost

A CatBoost classifier (version 1.1.1), a gradient boosting algorithm designed to handle tabular data,[30] was trained to classify cases into seven neuropathologic diagnostic categories using age at death, sex, brain weight, and 39 semi-quantitative gross findings. NA outputs from the extraction model were recoded as 0 before model training and prediction. The dataset was randomly split at the case level into a training set of 5,051 cases and a held-out test set of 562 cases. The CatBoost classifier was trained on the training set using 5,000 iterations, a learning rate of 0.01, a maximum tree depth of 8, an L2 leaf regularization coefficient of 8, and balanced class weights. Performance on the held-out test set was evaluated using sensitivity, specificity, accuracy, Cohen’s kappa, and receiver operating characteristic (ROC) curves with area under the curve (ROC-AUC).

### Text-based classification using an LLM

For the text-based classification approach, an additional text dataset was created to standardize the presentation of gross findings. Unmentioned findings, including those coded as NA by the extraction model, were represented by negative statements corresponding to a score of 0 in Supplementary Table S1, so that all 39 features were explicitly addressed for each case. This procedure treated unmentioned findings as absent; it did not distinguish undocumented abnormalities from true negative findings. Sentence order was randomized to reduce positional and template-related cues. Age at death, sex, and brain weight were also included in the input, as shown in Supplementary Table S2.

For text-based classification using an LLM, the GPT-4o-mini model (gpt-4o-mini-2024-07-18) was fine-tuned to classify each case into one of the seven neuropathologic diagnostic categories. The same training and held-out test sets used for score-based classification were used for fine-tuning and evaluation, respectively. The model was fine-tuned for three epochs with a batch size of 1 and a learning rate multiplier of 2, using the prompt provided in Supplementary Table S2. Performance was assessed using sensitivity, specificity, accuracy, and Cohen’s kappa. The LLM classifier returned diagnostic labels without continuous class scores; therefore, ROC curves and ROC-AUC values were not calculated for this model.

### Model interpretation

Global feature importance was calculated for the CatBoost model. Disease-specific feature contributions were visualized using SHapley Additive exPlanations (SHAP), which attributes model output to individual features based on game-theoretic principles.[22] Positive SHAP values supported the indicated diagnosis, whereas negative values opposed it. These contributions describe the fitted model and do not establish independent or causal associations between individual findings and disease.

### Statistical analysis

Descriptive statistics were used to summarize cohort characteristics and model performance. For frequency summaries, gross findings were classified as absent (score 0) or present (score ≥1), irrespective of severity (Supplementary Table S1). Categorical variables were compared using chi-square tests, and continuous variables were compared using the Kruskal-Wallis test. Statistical analyses and model training were performed using Python (version 3.9.15) and related libraries. A p value less than 0.05 was considered statistically significant.

## Results

### Clinicopathologic characteristics of the study cohort

Of 7,379 autopsy cases with available gross brain descriptions, 5,613 were included in the final cohort after exclusion of 1,766 cases in other or infrequent diagnostic categories (Fig. 1). The cohort comprised AD (n = 1,677), PSP (n = 1,450), LBD (n = 856), coexisting AD and LBD (AD-LBD; n = 746), MSA (n = 334), CBD (n = 280), and FTLD (n = 270). Overall, 54.8% of cases were male. The median age at death was 76 years, the median disease duration was 7 years, and the median brain weight was 1,100 g. AD had the highest median age at death at 80 years, whereas MSA had the lowest at 66 years. The proportion of male cases was highest in LBD at 73.0% and lowest in AD at 46.0%. Median brain weight was lowest in FTLD at 1,000 g and highest in LBD and MSA at 1,200 g. AD and AD-LBD had the highest median Braak neurofibrillary tangle stages and Thal phases. Among cases with available data, limbic TDP-43 pathology was most frequent in FTLD, whereas cerebrovascular disease was most frequent in AD. Some patients had more than one recorded clinical diagnosis. A matching clinical diagnosis was recorded in 88.4% of PSP cases and 84.4% of MSA cases. In contrast, pathologically confirmed CBD was clinically diagnosed as CBD in 45.7% and as PSP in 37.1% of cases. Among AD-LBD cases, the clinical diagnoses included AD in 51.6%, dementia with Lewy bodies in 27.9%, and Parkinson disease in 16.4% (Table 1).

**Table 1.**
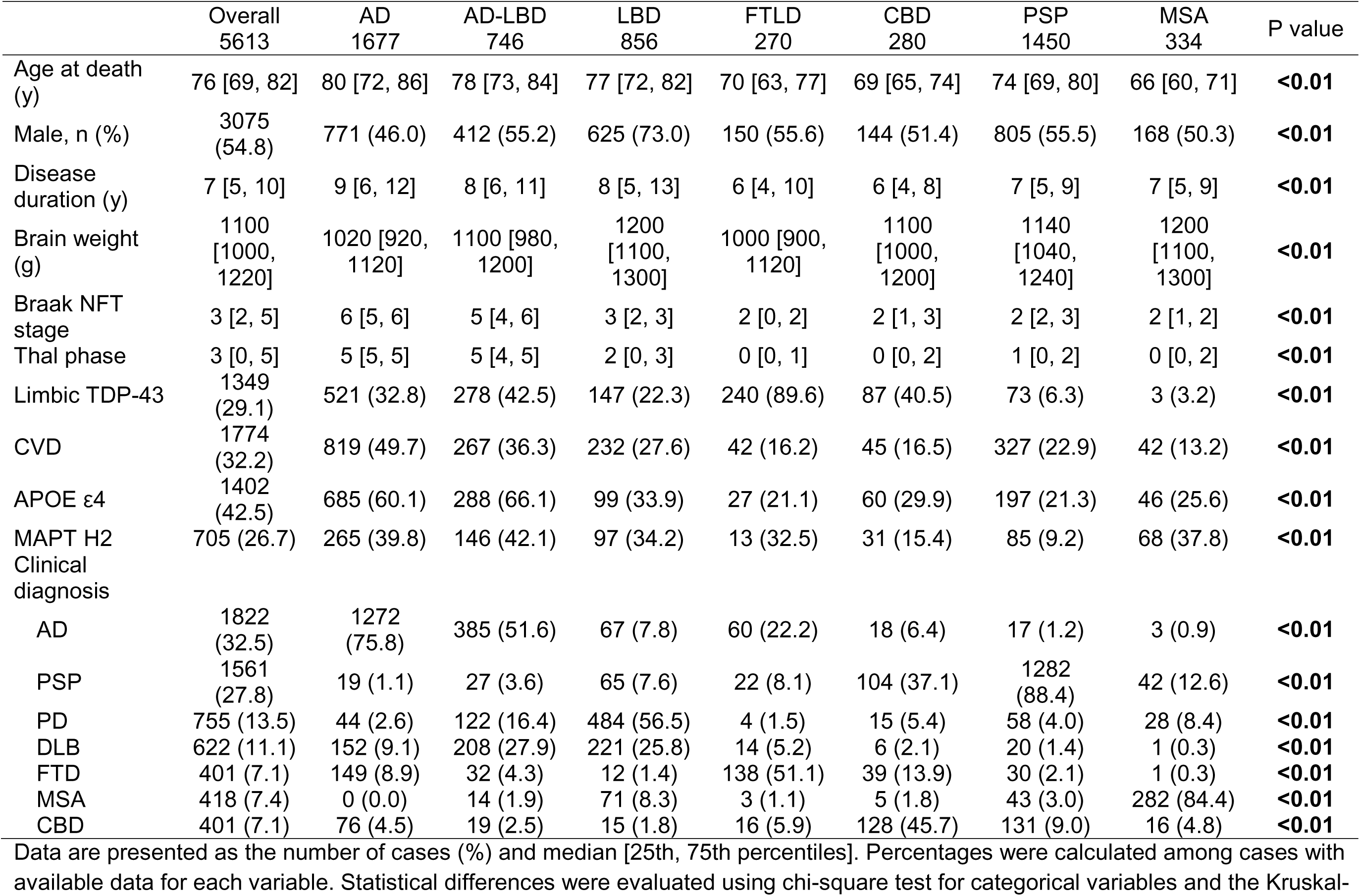
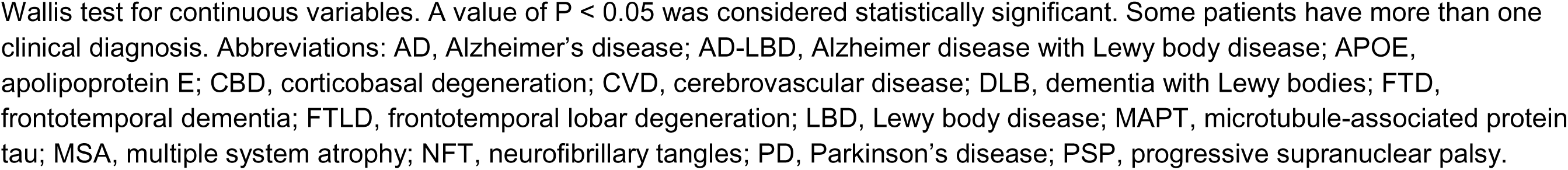
Clinicopathologic characteristics of patients.

|  | Overall<br>5613 | AD<br>1677 | AD-LBD<br>746 | LBD<br>856 | FTLD<br>270 | CBD<br>280 | PSP<br>1450 | MSA<br>334 | P value |
| --- | --- | --- | --- | --- | --- | --- | --- | --- | --- |
| Age at death (y) | 76 [69, 82] | 80 [72, 86] | 78 [73, 84] | 77 [72, 82] | 70 [63, 77] | 69 [65, 74] | 74 [69, 80] | 66 [60, 71] | **<0.01** |
| Male, n (%) | 3075 (54.8) | 771 (46.0) | 412 (55.2) | 625 (73.0) | 150 (55.6) | 144 (51.4) | 805 (55.5) | 168 (50.3) | **<0.01** |
| Disease duration (y) | 7 [5, 10] | 9 [6, 12] | 8 [6, 11] | 8 [5, 13] | 6 [4, 10] | 6 [4, 8] | 7 [5, 9] | 7 [5, 9] | **<0.01** |
| Brain weight (g) | 1100 [1000, 1220] | 1020 [920, 1120] | 1100 [980, 1200] | 1200 [1100, 1300] | 1000 [900, 1120] | 1100 [1000, 1200] | 1140 [1040, 1240] | 1200 [1100, 1300] | **<0.01** |
| Braak NFT stage | 3 [2, 5] | 6 [5, 6] | 5 [4, 6] | 3 [2, 3] | 2 [0, 2] | 2 [1, 3] | 2 [2, 3] | 2 [1, 2] | **<0.01** |
| Thal phase | 3 [0, 5] | 5 [5, 5] | 5 [4, 5] | 2 [0, 3] | 0 [0, 1] | 0 [0, 2] | 1 [0, 2] | 0 [0, 2] | **<0.01** |
| Limbic TDP-43 | 1349 (29.1) | 521 (32.8) | 278 (42.5) | 147 (22.3) | 240 (89.6) | 87 (40.5) | 73 (6.3) | 3 (3.2) | **<0.01** |
| CVD | 1774 (32.2) | 819 (49.7) | 267 (36.3) | 232 (27.6) | 42 (16.2) | 45 (16.5) | 327 (22.9) | 42 (13.2) | **<0.01** |
| APOE ε4 | 1402 (42.5) | 685 (60.1) | 288 (66.1) | 99 (33.9) | 27 (21.1) | 60 (29.9) | 197 (21.3) | 46 (25.6) | **<0.01** |
| MAPT H2 | 705 (26.7) | 265 (39.8) | 146 (42.1) | 97 (34.2) | 13 (32.5) | 31 (15.4) | 85 (9.2) | 68 (37.8) | **<0.01** |
| Clinical diagnosis |  |  |  |  |  |  |  |  |  |
| AD | 1822 (32.5) | 1272 (75.8) | 385 (51.6) | 67 (7.8) | 60 (22.2) | 18 (6.4) | 17 (1.2) | 3 (0.9) | **<0.01** |
| PSP | 1561 (27.8) | 19 (1.1) | 27 (3.6) | 65 (7.6) | 22 (8.1) | 104 (37.1) | 1282 (88.4) | 42 (12.6) | **<0.01** |
| PD | 755 (13.5) | 44 (2.6) | 122 (16.4) | 484 (56.5) | 4 (1.5) | 15 (5.4) | 58 (4.0) | 28 (8.4) | **<0.01** |
| DLB | 622 (11.1) | 152 (9.1) | 208 (27.9) | 221 (25.8) | 14 (5.2) | 6 (2.1) | 20 (1.4) | 1 (0.3) | **<0.01** |
| FTD | 401 (7.1) | 149 (8.9) | 32 (4.3) | 12 (1.4) | 138 (51.1) | 39 (13.9) | 30 (2.1) | 1 (0.3) | **<0.01** |
| MSA | 418 (7.4) | 0 (0.0) | 14 (1.9) | 71 (8.3) | 3 (1.1) | 5 (1.8) | 43 (3.0) | 282 (84.4) | **<0.01** |
| CBD | 401 (7.1) | 76 (4.5) | 19 (2.5) | 15 (1.8) | 16 (5.9) | 128 (45.7) | 131 (9.0) | 16 (4.8) | **<0.01** |

Data are presented as the number of cases (%) and median [25th, 75th percentiles]. Percentages were calculated among cases with available data for each variable. Statistical differences were evaluated using chi-square test for categorical variables and the Kruskal-
Wallis test for continuous variables. A value of $P < 0.05$ was considered statistically significant. Some patients have more than one clinical diagnosis. Abbreviations: AD, Alzheimer's disease; AD-LBD, Alzheimer disease with Lewy body disease; APOE, apolipoprotein E; CBD, corticobasal degeneration; CVD, cerebrovascular disease; DLB, dementia with Lewy bodies; FTD, frontotemporal dementia; FTLD, frontotemporal lobar degeneration; LBD, Lewy body disease; MAPT, microtubule-associated protein tau; MSA, multiple system atrophy; NFT, neurofibrillary tangles; PD, Parkinson's disease; PSP, progressive supranuclear palsy.

### Distribution of gross neuropathologic findings

The frequencies of 24 representative gross findings differed across diagnostic groups (Fig. 2). For visualization, these representative findings were selected from the 39 extracted features with consideration of anatomical overlap and feature importance. The full set of parameters is detailed in Supplementary Table S3. AD frequently showed temporal lobe atrophy (76%), hippocampal atrophy (75%), and cerebral white matter abnormalities (60%). AD-LBD showed intermediate frequencies of temporal lobe atrophy (56%) and hippocampal atrophy (68%). In LBD, substantia nigra and locus coeruleus depigmentation were frequent at 89% and 87%, respectively, whereas temporal lobe and hippocampal atrophy were less frequent at 10% and 29%.

**Fig. 2.**
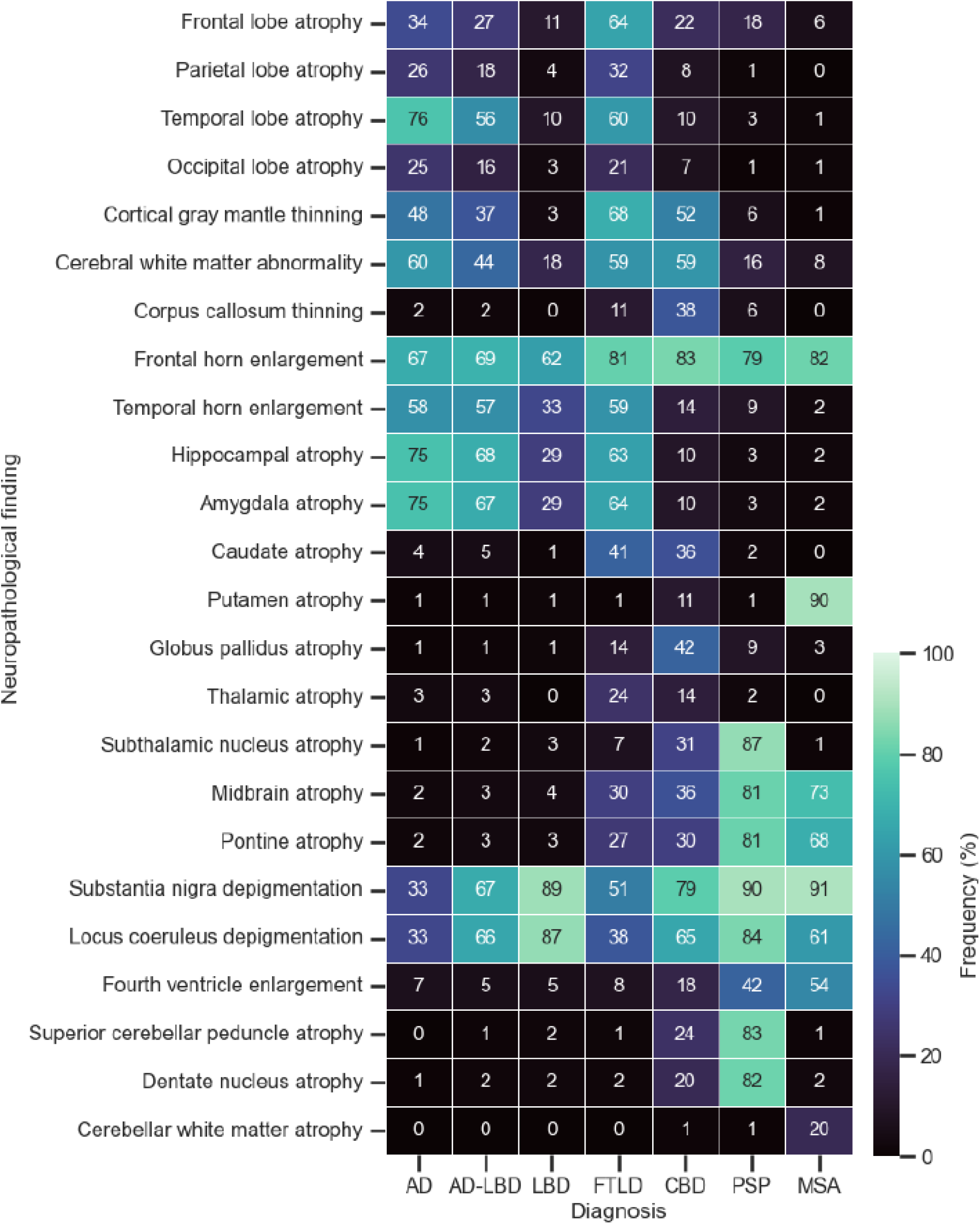
Gross neuropathological findings across neuropathologic diagnoses. The heatmap shows the frequency (%) of each gross neuropathological finding across the seven neuropathologic diagnostic groups. Values in each cell indicate the frequency (%) of the corresponding finding. Abbreviations: AD, Alzheimer disease; AD-LBD, Alzheimer disease with Lewy body disease; CBD, corticobasal degeneration; FTLD, frontotemporal lobar degeneration; LBD, Lewy body disease; MSA, multiple system atrophy; PSP, progressive supranuclear palsy.

FTLD frequently showed frontal lobe atrophy (64%), temporal lobe atrophy (60%), cortical gray mantle thinning (68%), and caudate atrophy (41%). CBD showed corpus callosum thinning in 38%, caudate atrophy in 36%, globus pallidus atrophy in 42%, subthalamic nucleus atrophy in 31%, and midbrain atrophy in 36%. PSP had high frequencies of subthalamic nucleus atrophy (87%), midbrain atrophy (81%), superior cerebellar peduncle atrophy (83%), pontine atrophy (81%), and dentate nucleus atrophy (82%). MSA was characterized by putamen atrophy (90%), midbrain atrophy (73%), and pontine atrophy (68%). Substantia nigra and locus coeruleus depigmentation were common across several parkinsonian disorders and were not confined to a single diagnosis.

### Diagnostic performance on the held-out test set

The held-out test dataset comprised 562 cases: AD (n = 167), AD-LBD (n = 70), LBD (n = 94), FTLD (n = 19), CBD (n = 35), PSP (n = 138), and MSA (n = 39). The score-based CatBoost model achieved an overall accuracy of 0.73 and Cohen’s kappa of 0.65 (**Table 2**). The micro-, macro-, and weighted ROC-AUC values were 0.95, 0.92, and 0.93, respectively. Class-specific ROC-AUC values were highest for PSP and MSA (0.99 for both), followed by LBD (0.94), FTLD (0.93), AD (0.91), CBD (0.89), and AD-LBD (0.78) (**Fig. 3a**). Ten-fold cross-validation reproduced the diagnostic pattern observed in the held-out test analysis (**Supplementary Fig. S1**).

**Table 2.**
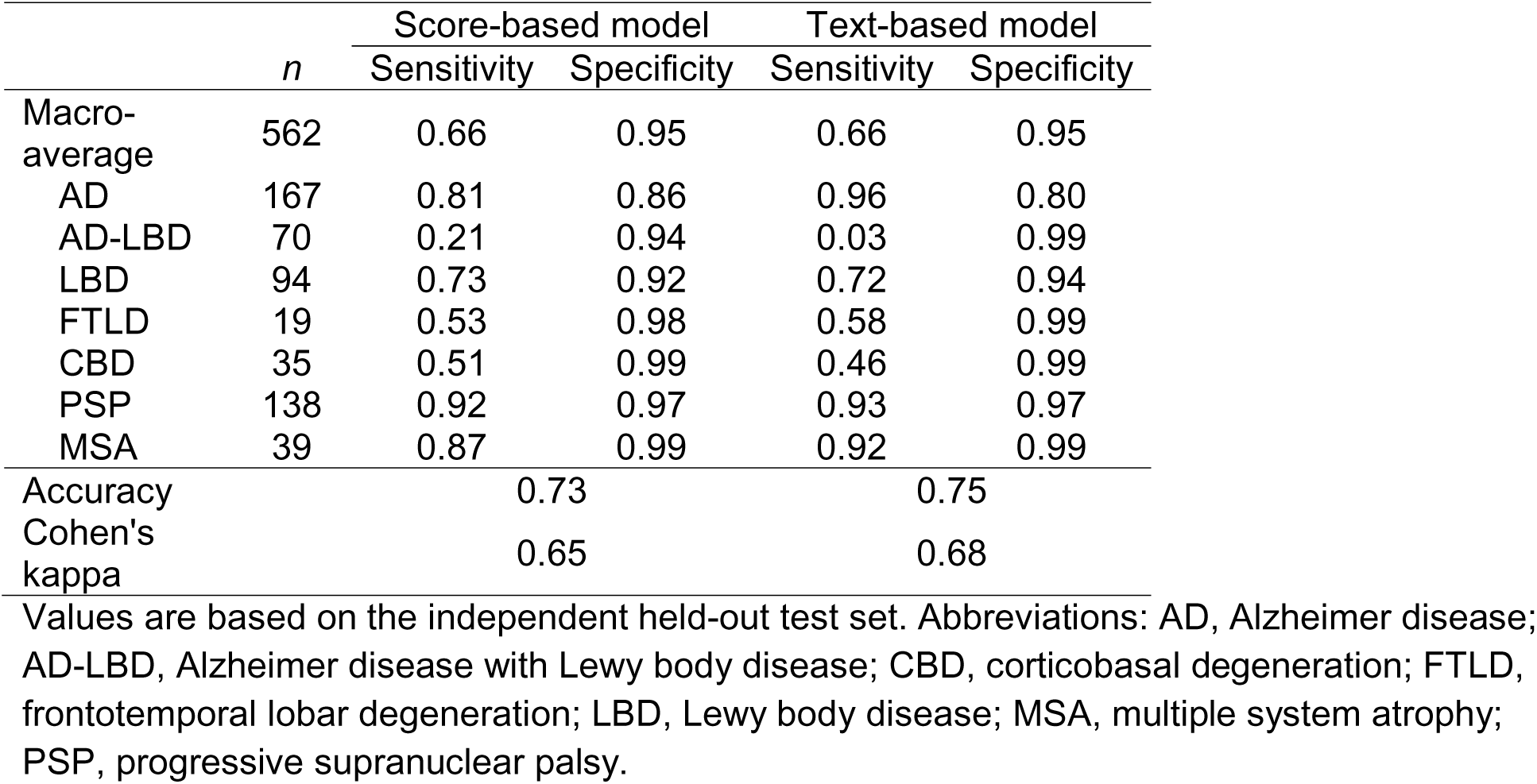
Diagnostic performance on the held-out test set.

|  | *n* | Score-based model |  | Text-based model |  |
| --- | --- | --- | --- | --- | --- |
|  |  | Sensitivity | Specificity | Sensitivity | Specificity |
| Macro-average | 562 | 0.66 | 0.95 | 0.66 | 0.95 |
| AD | 167 | 0.81 | 0.86 | 0.96 | 0.80 |
| AD-LBD | 70 | 0.21 | 0.94 | 0.03 | 0.99 |
| LBD | 94 | 0.73 | 0.92 | 0.72 | 0.94 |
| FTLD | 19 | 0.53 | 0.98 | 0.58 | 0.99 |
| CBD | 35 | 0.51 | 0.99 | 0.46 | 0.99 |
| PSP | 138 | 0.92 | 0.97 | 0.93 | 0.97 |
| MSA | 39 | 0.87 | 0.99 | 0.92 | 0.99 |
| Accuracy |  | 0.73 |  | 0.75 |  |
| Cohen's kappa |  | 0.65 |  | 0.68 |  |

Values are based on the independent held-out test set. Abbreviations: AD, Alzheimer disease; AD-LBD, Alzheimer disease with Lewy body disease; CBD, corticobasal degeneration; FTLD, frontotemporal lobar degeneration; LBD, Lewy body disease; MSA, multiple system atrophy; PSP, progressive supranuclear palsy.

**Fig. 3.**
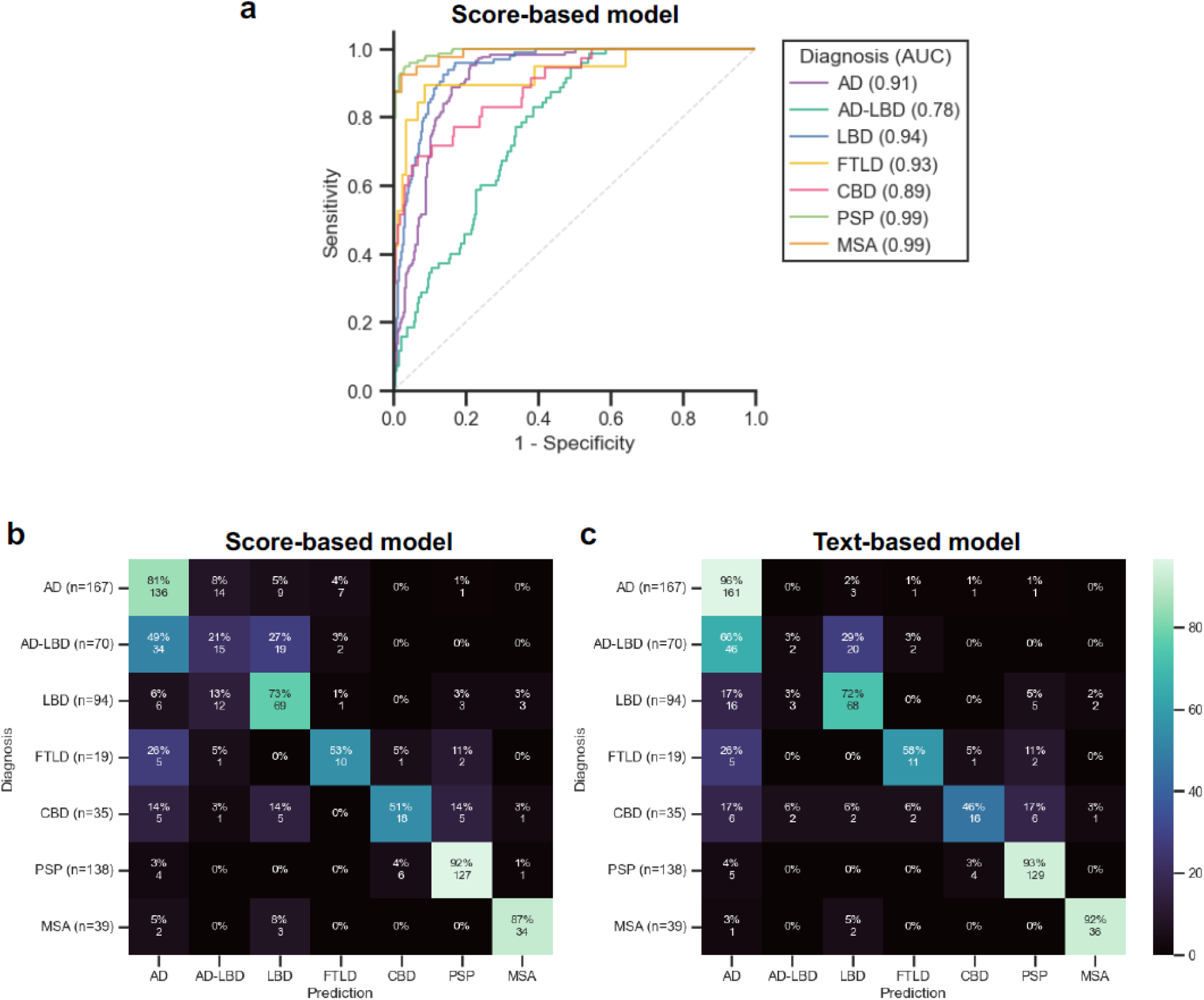
Diagnostic performance of score-based and text-based classification models in the held-out test dataset. **a** Receiver operating characteristic curves for the score-based CatBoost model, with micro-, macro-, and weighted-average ROC-AUCs of 0.95, 0.92, and 0.93, respectively. **b, c** Confusion matrices show diagnostic classification using the score-based CatBoost model and the text-based LLM (GPT-4o-mini) model, respectively. Rows indicate the final neuropathologic diagnoses, and columns indicate the predicted diagnoses. Performance metrics are presented in Table 2. Abbreviations: AD, Alzheimer disease; AD-LBD, Alzheimer disease with Lewy body disease; AUC, area under the curve; CBD, corticobasal degeneration; FTLD, frontotemporal lobar degeneration; LBD, Lewy body disease; MSA, multiple system atrophy; PSP, progressive supranuclear palsy.

On the same test set, the fine-tuned LLM-based classifier achieved an overall accuracy of 0.75 and Cohen’s kappa of 0.68 (Table 2). Compared with CatBoost, the LLM-based classifier showed higher sensitivity for AD and MSA, and lower sensitivity for CBD and AD-LBD. Continuous class scores were unavailable for the LLM classifier, so ROC analyses were performed only for CatBoost.

### Disease-specific performance and error patterns

Both classifiers showed high sensitivity for PSP and MSA (Table 2 and Figs. 3b and c). For PSP, sensitivity was 0.92 with CatBoost and 0.93 with the LLM classifier. For MSA, the corresponding values were 0.87 and 0.92. Sensitivities for AD were 0.81 and 0.96, and those for LBD were 0.73 and 0.72. FTLD and CBD showed lower sensitivities. CatBoost sensitivities were 0.53 for FTLD and 0.51 for CBD, whereas LLM sensitivities were 0.58 and 0.46, respectively.

AD-LBD had the lowest sensitivity with both classifiers. CatBoost correctly classified 15 of 70 cases (21%), whereas the LLM classifier correctly classified 2 of 70 cases (3%). CatBoost classified 49% of AD-LBD cases as AD and 27% as LBD. The LLM classifier classified 66% as AD and 29% as LBD. Thus, most classification errors for AD-LBD involved one of its two component pathologies. The LLM classifier had slightly higher overall accuracy (0.75 vs 0.73), whereas its macro-average sensitivity was comparable to that of CatBoost (both 0.66). These findings do not indicate uniform superiority of either classifier across diagnostic groups.

### Feature importance and SHAP interpretation in the table-based model

Global feature importance analysis in the CatBoost model identified age (10.40) and brain weight (9.31) as the two highest-ranked variables (Supplementary Table S3). Among macroscopic findings, the highest-ranked features were substantia nigra depigmentation (4.51), cortical atrophy (4.27), and locus coeruleus depigmentation (4.23).

Disease-specific SHAP analyses identified distinct patterns of feature contributions for PSP and MSA. Subthalamic nucleus atrophy was the strongest positive contributor to PSP prediction. Superior cerebellar peduncle atrophy, dentate hilus discoloration, dentate nucleus atrophy, and midbrain atrophy also supported PSP prediction (Fig. 4).

**Fig. 4.**
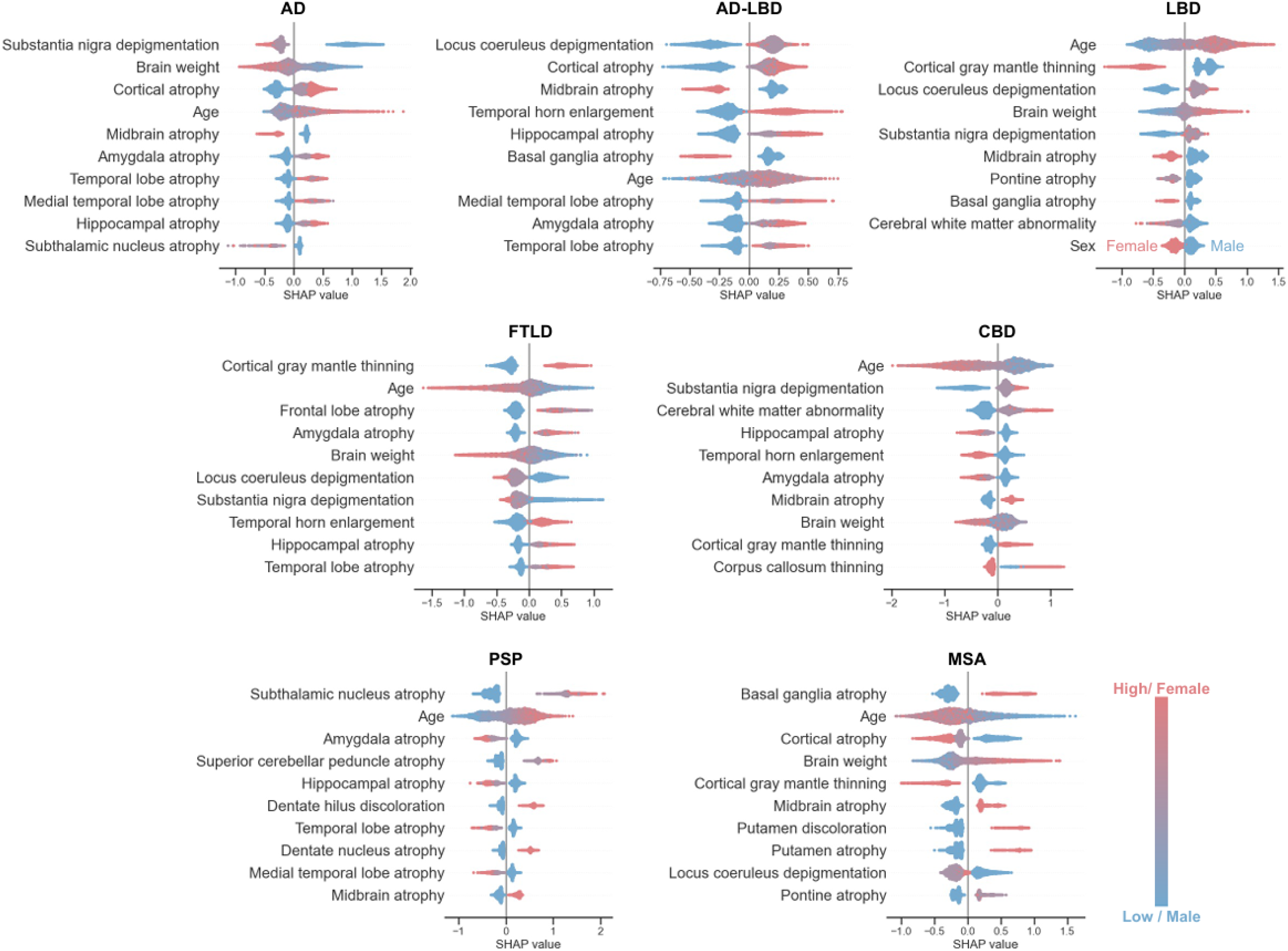
SHAP analysis in the CatBoost score-based model. Plots displaying SHAP (SHapley Additive exPlanations) values for the top 10 features across neuropathological diagnoses. Each dot represents one case. Positive SHAP values support the indicated diagnosis, whereas negative values oppose it. Dot color indicates feature value, with blue representing lower values and red representing higher values. Abbreviations: AD, Alzheimer disease; AD-LBD, Alzheimer disease with Lewy body disease; CBD, corticobasal degeneration; FTLD, frontotemporal lobar degeneration; LBD, Lewy body disease; MSA, multiple system atrophy; PSP, progressive supranuclear palsy.

For MSA, basal ganglia atrophy, putamen atrophy, and putamen discoloration supported prediction. Other supportive features included the absence of cortical atrophy, higher brain weight, and pontine atrophy. These contributions were consistent with the regional distribution of degeneration in MSA (Fig. 4).

Cortical and limbic findings contributed to AD, FTLD, and CBD predictions. For AD, cortical, amygdala, temporal lobe, and hippocampal atrophy supported prediction. Preserved nigral pigmentation and the absence of midbrain atrophy also supported AD predictions. For FTLD, cortical gray mantle thinning, frontal and temporal lobe atrophy, and amygdala and hippocampal atrophy contributed positively. For CBD, supportive findings included substantia nigra depigmentation, cerebral white matter abnormalities, midbrain atrophy, cortical gray mantle thinning, and corpus callosum thinning (Fig. 4).

For LBD, locus coeruleus and substantia nigra depigmentation supported prediction. Features contributing to AD-LBD predictions included cortical and medial temporal atrophy and locus coeruleus depigmentation. Despite these contributions, the feature set and classification approach did not reliably distinguish AD-LBD from its component pathologies (Fig. 4).

## Discussion

In this large autopsy-based study, we tested whether macroscopic brain findings described in neuropathology reports could predict final neuropathologic diagnoses of major neurodegenerative diseases. Using 5,613 autopsy-confirmed cases from the Mayo Clinic Brain Bank, narrative gross descriptions were converted into semi-quantitative variables with a fine-tuned LLM and evaluated with a score-based CatBoost classifier and a text-based LLM classifier. Models based on gross-description-derived variables, together with age, sex, and brain weight, showed substantial variation in diagnostic performance across groups. PSP and MSA were accurately classified, whereas AD-LBD was the least accurately predicted. SHAP analysis showed that disease-specific gross features, including subthalamic nucleus atrophy in PSP and putamen atrophy or discoloration in MSA, were among the most informative predictors. These results provide quantitative support for long-standing gross neuropathology teaching and identify anatomical features for further imaging-pathology correlation.

The high sensitivity for PSP and MSA is consistent with their characteristic gross pathologic patterns. PSP commonly involves the subthalamic nucleus, midbrain, superior cerebellar peduncle, and dentate nucleus, often with substantia nigra pigment loss.[5] MSA produces recognizable involvement of the putamen, pons, cerebellum, and cerebellar white matter.[14, 35] In contrast, the combined AD-LBD category was poorly predicted by both models. The extent and distribution of AD and Lewy body pathology vary, and the two components may contribute differently to the recorded gross phenotype.[25] Mixed neurodegenerative pathologies are common in aging brains and complicate clinicopathologic classification.[7, 31, 34] However, the low sensitivity for AD-LBD should not be interpreted as evidence that mixed pathology lacks macroscopic correlates. The available features, the representation of combined pathology as a single mutually exclusive class, and class imbalance may also have limited discrimination. Future studies could evaluate multilabel approaches that predict AD and LBD components separately.

The lower sensitivity for CBD warrants consideration of the information available to the models. Corpus callosum thinning and cortical and subcortical abnormalities contributed to CBD prediction, but the feature set did not encode hemispheric asymmetry. Because one hemisphere was frozen and the other was fixed for neuropathologic assessment, systematic bilateral comparison of regional atrophy in fixed tissue was not possible. This limitation may have reduced the representation of the asymmetric cortical pattern associated with CBD. Lower sensitivity therefore reflects the performance of the recorded features under this protocol and does not establish that gross examination has little discriminatory value for CBD.

The two modeling approaches showed different patterns of performance. CatBoost provided continuous class scores for ROC analysis and supported feature attribution through SHAP. The LLM classifier had slightly higher overall accuracy but markedly lower sensitivity for AD-LBD, while macro-average sensitivity was similar. These findings do not establish uniform superiority of either model. Differences in the learning objectives and handling of class imbalance may also have contributed, particularly because balanced class weights were used for CatBoost. The models should therefore be compared using both overall and class-specific performance.

Our approach extends brain bank studies that used LLMs to structure narrative records.[18, 24] Stroganov et al. extracted macroscopic and microscopic observations from NIH NeuroBioBank reports.[33] Ono et al. used fine-tuned language models to extract longitudinal clinical information for PSP subtype analysis.[28] A subsequent study applied this approach to neuropathologic prediction in parkinsonism.[29] The present study focuses on gross neuropathology descriptions and the diagnostic information contained in structured macroscopic observations.

The present analysis was based on report text rather than primary gross images. This design has practical value because historical brain bank archives contain decades of narrative autopsy reports that can be analyzed retrospectively. At the same time, report-based analysis depends on reporting style, terminology, and the level of detail recorded by the neuropathologist. Recent reviews have emphasized the potential of computational neuropathology and machine learning for neurodegenerative disease research, while also noting barriers to image-based methods, including limited expert annotations, institutional variability, access to large-scale digitized images, and challenges in data sharing.[12] Report-based analysis therefore provides a complementary approach that can make use of existing brain bank archives. Future studies could incorporate standardized gross photographs and antemortem imaging to test whether multimodal models can learn visual correlates of disease-specific gross pathology directly.

These results suggest potential applications in neuropathology education. Disease-specific SHAP plots can illustrate how combinations of recorded gross findings contribute to model predictions. Educational benefit and effects on tissue sampling or immunohistochemical workup were not evaluated in this study. Such applications would require external validation and prospective assessment. The classifier was trained to choose among seven selected diagnostic categories and should not be interpreted as evaluating the full differential diagnosis of an unselected autopsy case.

Several informative gross findings correspond to previously reported MRI correlates. Fujioka et al. reported reduced subthalamic nucleus volumes in a small clinical MRI study of PSP, supporting further evaluation of this structure rather than establishing a definitive imaging marker.[8] In an autopsy-confirmed MRI study, Illán-Gala et al. described greater perirolandic, putaminal, and corpus callosum atrophy in CBD than in PSP, whereas PSP showed greater brainstem involvement.[10] The corpus callosum findings provide a relevant comparison with our gross observations in CBD. However, these findings do not directly validate caudate atrophy as an MRI discriminator between CBD and PSP.

Caudate atrophy was more frequent in CBD than in PSP in our cohort. Caudate atrophy was also common in the heterogeneous FTLD group. Previous studies have reported caudate involvement in FTLD-TDP type C and FTLD-FUS.[2, 11, 13] The contributions of individual molecular subtypes to the findings in our cohort were not evaluated. Our previous study identified caudate tau-positive threads as an important histologic discriminator between CBD and PSP.[20] This supports the relevance of the region but does not demonstrate a direct relationship between tau burden and gross atrophy. No direct MRI-pathology comparison was performed in the present study. Caudate atrophy should therefore be evaluated as a candidate feature within the differential diagnosis of CBD and PSP, rather than as a CBD-specific imaging marker.

This study has several limitations. First, it was conducted at a single brain bank, and the findings may reflect local reporting practices, terminology, and sampling procedures. Second, the protocol of freezing one hemisphere and fixing the other precluded systematic bilateral assessment of regional atrophy in fixed tissue. Hemispheric asymmetry was therefore not represented in the model features, which may have limited classification of CBD. Third, NA outputs were treated as absent in both classifiers. This approach could not distinguish true negative findings from undocumented abnormalities and may limit generalizability to reports with different levels of detail. Fourth, extraction errors could propagate to diagnostic classification, and high overall extraction agreement does not establish equivalent performance for every feature. Fifth, diagnostic groups were imbalanced, and performance estimates for smaller test-set groups require cautious interpretation. The incremental value of the 39 gross findings beyond age, sex, and brain weight was not separately evaluated. Finally, the cohort comprised seven selected neuropathologic categories. External validation is needed before extending the results to other brain banks or to unselected autopsy populations.

Despite these limitations, our findings show that narrative gross brain descriptions contain diagnostic information that can be structured by an LLM and used for machine learning-based diagnostic classification. The models performed best for disorders with distinctive recorded gross patterns, whereas the combined AD-LBD category remained difficult to classify. By combining LLM-assisted extraction, table-based classification, and SHAP analysis, this study provides a framework for quantitative analysis of recorded gross neuropathology. Future studies should validate this approach externally and evaluate multimodal models incorporating standardized gross photography, antemortem imaging, and report text.

## Acknowledgments

We thank the patients and their families who donated brains to support neurodegenerative disease research. We also thank the Mayo Clinic Brain Bank and neuropathology laboratory staff for tissue processing, histology, immunohistochemistry, data management, and coordination of brain donations.

## Author Contributions

D.O. and S.K. contributed to conceptualization and study design. D.O. led the formal analysis, machine learning methodology, validation, visualization, and interpretation of model outputs, and contributed to review and editing of the manuscript. D.W.D. contributed to case resources and neuropathologic diagnoses and critically reviewed the manuscript. S.K. contributed to data curation, interpretation of neuropathologic findings, and writing of the original draft. All authors approved the final version.

## Data Availability

The CatBoost model trained in this study will be made publicly available at https://github.com/neurologydatascience/grossgnosis upon publication. The de-identified data that support the findings of this study may be available from the corresponding author upon reasonable request and subject to institutional policies and applicable data sharing agreements.

## References

1 Bigio EH (2013) Making the diagnosis of frontotemporal lobar degeneration. Arch Pathol Lab Med 137: 314–325 10.5858/arpa.2012-0075-RA

2 Bocchetta M, Iglesias Espinosa MDM, Lashley T, Warren JD, Rohrer JD (2020) In vivo staging of frontotemporal lobar degeneration TDP-43 type C pathology. Alzheimers Res Ther 12: 34 10.1186/s13195-020-00600-x

3 Cairns NJ, Bigio EH, Mackenzie IR, Neumann M, Lee VM, Hatanpaa KJ et al (2007) Neuropathologic diagnostic and nosologic criteria for frontotemporal lobar degeneration: consensus of the Consortium for Frontotemporal Lobar Degeneration. Acta Neuropathol 114: 5–22 10.1007/s00401-007-0237-2

4 Dickson DW, Bergeron C, Chin SS, Duyckaerts C, Horoupian D, Ikeda K et al (2002) Office of Rare Diseases neuropathologic criteria for corticobasal degeneration. J Neuropathol Exp Neurol 61: 935–946 10.1093/jnen/61.11.935

5 Dickson DW, Rademakers R, Hutton ML (2007) Progressive supranuclear palsy: pathology and genetics. Brain Pathol 17: 74–82 10.1111/j.1750-3639.2007.00054.x

6 Dugger BN, Dickson DW (2017) Pathology of Neurodegenerative Diseases. Cold Spring Harb Perspect Biol 9: a028035 10.1101/cshperspect.a028035

7 Forrest SL, Kovacs GG (2023) Current Concepts of Mixed Pathologies in Neurodegenerative Diseases. Can J Neurol Sci 50: 329–345 10.1017/cjn.2022.34

8 Fujioka S, Morishita T, Takano K, Takahashi N, Kurihara K, Nishida A et al (2021) A novel diagnostic marker for progressive supranuclear palsy targeting atrophy of the subthalamic nucleus. J Neurol Sci 423: 117366 10.1016/j.jns.2021.117366

9 Hashizume Y (2022) Macroscopic findings of brain with dementia. Neuropathology 42: 353–366 10.1111/neup.12785

10 Illan-Gala I, Nigro S, VandeVrede L, Falgas N, Heuer HW, Painous C et al (2022) Diagnostic Accuracy of Magnetic Resonance Imaging Measures of Brain Atrophy Across the Spectrum of Progressive Supranuclear Palsy and Corticobasal Degeneration. JAMA Netw Open 5: e229588 10.1001/jamanetworkopen.2022.9588

11 Josephs KA, Whitwell JL, Parisi JE, Petersen RC, Boeve BF, Jack CR, Jr. et al (2010) Caudate atrophy on MRI is a characteristic feature of FTLD-FUS. Eur J Neurol 17: 969–975 10.1111/j.1468-1331.2010.02975.x

12 Julian DR, Bahramy A, Neal M, Pearce TM, Kofler J (2025) Current Advancements in Digital Neuropathology and Machine Learning for the Study of Neurodegenerative Diseases. Am J Pathol 195: 2102–2117 10.1016/j.ajpath.2024.12.018

13 Kawles A, Nishihira Y, Feldman A, Gill N, Minogue G, Keszycki R et al (2022) Cortical and subcortical pathological burden and neuronal loss in an autopsy series of FTLD-TDP-type C. Brain 145: 1069–1078 10.1093/brain/awab368

14 Koga S, Dickson DW (2018) Recent advances in neuropathology, biomarkers and therapeutic approach of multiple system atrophy. J Neurol Neurosurg Psychiatry 89: 175–184 10.1136/jnnp-2017-315813

15 Koga S, Josephs KA, Aiba I, Yoshida M, Dickson DW (2022) Neuropathology and emerging biomarkers in corticobasal syndrome. J Neurol Neurosurg Psychiatry 93: 919–929 10.1136/jnnp-2021-328586

16 Koga S, Kouri N, Walton RL, Ebbert MTW, Josephs KA, Litvan I et al (2018) Corticobasal degeneration with TDP-43 pathology presenting with progressive supranuclear palsy syndrome: a distinct clinicopathologic subtype. Acta Neuropathol 136: 389–404 10.1007/s00401-018-1878-z

17 Koga S, Li F, Zhao N, Roemer SF, Ferman TJ, Wernick AI et al (2020) Clinicopathologic and genetic features of multiple system atrophy with Lewy body disease. Brain Pathol 30: 766–778 10.1111/bpa.12839

18 Koga S, Martin NB, Dickson DW (2023) Evaluating the performance of large language models: ChatGPT and Google Bard in generating differential diagnoses in clinicopathological conferences of neurodegenerative disorders. Brain Pathol: e13207 10.1111/bpa.13207

19 Koga S, Roemer SF, Tipton PW, Low PA, Josephs KA, Dickson DW (2020) Cerebrovascular pathology and misdiagnosis of multiple system atrophy: An autopsy study. Parkinsonism Relat Disord 75: 34–40 10.1016/j.parkreldis.2020.05.018

20 Koga S, Zhou X, Dickson DW (2021) Machine learning-based decision tree classifier for the diagnosis of progressive supranuclear palsy and corticobasal degeneration. Neuropathol Appl Neurobiol 47: 931–941 10.1111/nan.12710

21 Li K, Rashid T, Li J, Honnorat N, Nirmala AB, Fadaee E et al (2023) Postmortem Brain Imaging in Alzheimer’s Disease and Related Dementias: The South Texas Alzheimer’s Disease Research Center Repository. J Alzheimers Dis 96: 1267–1283 10.3233/JAD-230389

22 Lundberg SM, Lee S-I (2017) A Unified Approach to Interpreting Model Predictions. In: Guyon I, Luxburg UV, Bengio S, Wallach H, Fergus R, Vishwanathan S, Garnett R (eds) Advances in Neural Information Processing Systems. Curran Associates, Inc., pp 4765–4774

23 Mackenzie IR, Neumann M, Bigio EH, Cairns NJ, Alafuzoff I, Kril J et al (2010) Nomenclature and nosology for neuropathologic subtypes of frontotemporal lobar degeneration: an update. Acta Neuropathol 119: 1–4 10.1007/s00401-009-0612-2

24 Martin NB, Sekiya H, Kim M, Dickson DW, Koga S (2023) Brain Bank Questionnaire Helps in Differential Diagnosis of Movement Disorders: An Autopsy Study of 150 Patients. Mov Disord Clin Pract 10: 1131–1135 10.1002/mdc3.13788

25 McKeith IG, Boeve BF, Dickson DW, Halliday G, Taylor JP, Weintraub D et al (2017) Diagnosis and management of dementia with Lewy bodies: Fourth consensus report of the DLB Consortium. Neurology 89: 88–100 10.1212/WNL.0000000000004058

26 Montine TJ, Phelps CH, Beach TG, Bigio EH, Cairns NJ, Dickson DW et al (2012) National Institute on Aging-Alzheimer’s Association guidelines for the neuropathologic assessment of Alzheimer’s disease: a practical approach. Acta Neuropathol 123: 1–11 10.1007/s00401-011-0910-3

27 Murakami A, Koga S, Sekiya H, Nakamura M, Yakushiji Y, Dickson DW (2025) Frontotemporal Lobar degeneration with TDP-43 presenting as progressive supranuclear palsy syndrome. Acta Neuropathol Commun 13: 151 10.1186/s40478-025-02058-0

28 Ono D, Sekiya H, Ghayal NB, Maier AR, Roemer SF, Uitti RJ et al (2025) Early subtypes and progressions of progressive supranuclear palsy: a data-driven brain bank study. J Neurol 272: 704 10.1007/s00415-025-13459-5

29 Ono D, Sekiya H, Maier AR, Graff-Radford NR, Wszolek ZK, Dickson DW (2026) Chronological Diagnostic Algorithm Predicting Neuropathology in Parkinsonism. Ann Neurol 99: 1405–1414 10.1002/ana.78193

30 Prokhorenkova L, Gusev G, Vorobev A, Dorogush AV, Gulin A (2018) CatBoost: unbiased boosting with categorical features. In: Bengio S, Wallach H, Larochelle H, Grauman K, Cesa-Bianchi N, Garnett R (eds) Advances in Neural Information Processing Systems. Curran Associates, Inc., pp 6638–6648

31 Robinson JL, Lee EB, Xie SX, Rennert L, Suh E, Bredenberg C et al (2018) Neurodegenerative disease concomitant proteinopathies are prevalent, age-related and APOE4-associated. Brain 141: 2181–2193 10.1093/brain/awy146

32 Sekiya H, Koga S, Murakami A, DeTure M, Ross OA, Uitti RJ et al (2024) Frequency of Comorbid Pathologies and Their Clinical Impact in Multiple System Atrophy. Mov Disord 39: 380–390 10.1002/mds.29670

33 Stroganov O, Schedlbauer A, Lorenzen E, Kadhim A, Lobanova A, Lewis DA, et al (2024) Unpacking unstructured data: A pilot study on extracting insights from neuropathological reports of Parkinson’s Disease patients using large language models. Biol Methods Protoc 9: bpae072 10.1093/biomethods/bpae072

34 Toledo JB, Abdelnour C, Weil RS, Ferreira D, Rodriguez-Porcel F, Pilotto A et al (2023) Dementia with Lewy bodies: Impact of co-pathologies and implications for clinical trial design. Alzheimers Dement 19: 318–332 10.1002/alz.12814

35 Trojanowski JQ, Revesz T, Neuropathology Working Group on MSA (2007) Proposed neuropathological criteria for the post mortem diagnosis of multiple system atrophy. Neuropathol Appl Neurobiol 33: 615–620 10.1111/j.1365-2990.2007.00907.x

36 Tunold JA, Tan MMX, Koga S, Geut H, Rozemuller AJM, Valentino R et al (2023) Lysosomal polygenic risk is associated with the severity of neuropathology in Lewy body disease. Brain 146: 4077–4087 10.1093/brain/awad183

37 Walton RL, Koga S, Beasley AI, White LJ, Griesacker T, Murray ME et al (2024) Role of GBA variants in Lewy body disease neuropathology. Acta Neuropathol 147: 54 10.1007/s00401-024-02699-w

