## Supplementary information for "Diagnostic Value of Large Language Model-Extracted Gross Brain Findings in Neurodegenerative Diseases"

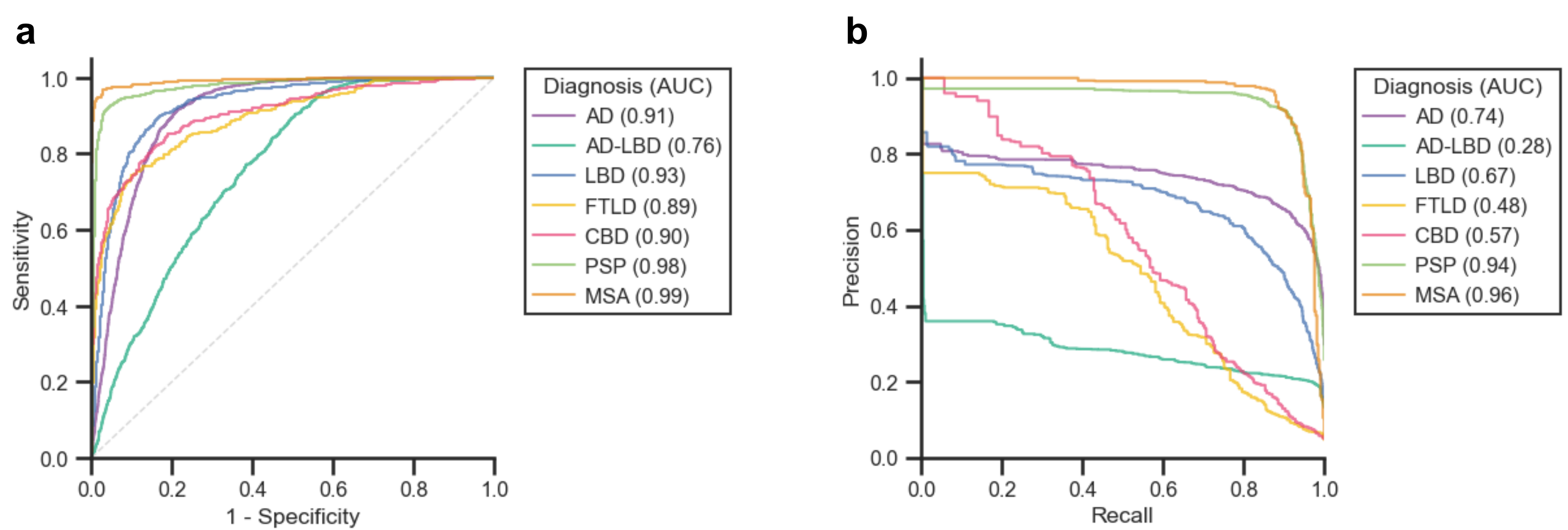

**Supplementary Fig. S1**. Diagnostic performance of the CatBoost model assessed using 10-fold cross-validation. **a** Receiver operating characteristic curves and **b** precision–recall curves for seven neuropathological diagnostic labels are shown. The area under the curve (AUC) for each diagnostic label is indicated in the legend. PSP and MSA showed the highest diagnostic performance, whereas AD-LBD showed the lowest performance. Abbreviations: AD, Alzheimer disease; AD-LBD, combined Alzheimer disease and Lewy body disease; CBD, corticobasal degeneration; FTLD, frontotemporal lobar degeneration; LBD, Lewy body disease; MSA, multiple system atrophy; PSP, progressive supranuclear palsy.

**Supplementary Table S1** Scoring Criteria and Example Descriptions for Macroscopic Findings

| Findings | Score | Example descriptions |
| --- | --- | --- |
| Amygdala atrophy | 0 | The amygdala has no atrophy |
|  | 1 | The amygdala is slightly atrophic |
|  | 2 | The amygdala is atrophic |
|  | 3 | The amygdala is markedly atrophic |
| Anterior commissure abnormality | 0 | The anterior commissure is unremarkable |
|  | 1 | The anterior commissure is thin and discolored |
| Anterior cortical atrophy | 0 | The sulci and gyri reveal no anterior cortical atrophy |
|  | 1 | The sulci and gyri reveal anterior cortical atrophy |
|  | 2 | The sulci and gyri reveal marked anterior cortical atrophy, |
| Atherosclerosis | 0 | The available blood vessels at the base of the brain have no atherosclerosis |
|  | 1 | The available blood vessels at the base of the brain show minimal atherosclerosis |
|  | 2 | The available blood vessels at the base of the brain show atherosclerosis |
|  | 3 | The available blood vessels at the base of the brain show marked atherosclerosis |
| Basal ganglia atrophy | 0 | The basal ganglia are unremarkable |
|  | 1 | The basal ganglia show atrophy |
|  | 2 | The basal ganglia show marked atrophy |
| Caudate atrophy | 0 | The caudate nucleus is unremarkable |
|  | 1 | The caudate nucleus shows atrophy |
|  | 2 | The caudate nucleus shows marked atrophy |
| Cerebellar white matter atrophy | 0 | The cerebellar white matter is intact |
|  | 1 | The cerebellar white matter has atrophy |
|  | 2 | The cerebellar white matter is markedly atrophic |
| Cerebral white matter abnormality | 0 | The subjacent white matter shows no unusual features |
|  | 1 | The subjacent white matter shows attenuation and discoloration |
|  | 2 | The subjacent white matter shows marked attenuation and discoloration |
| Corpus callosum thinning | 0 | The corpus callosum has no atrophy |
|  | 1 | The corpus callosum is thin and discolored |
| Cortical atrophy | 0 | The sulci and gyri reveal no cortical atrophy over the convexity |
|  | 1 | The sulci and gyri reveal cortical atrophy over the convexity |
|  | 2 | The sulci and gyri reveal marked cortical atrophy over the convexity |
| Cortical gray mantle thinning | 0 | The cortical gray mantle is normal in thickness |
|  | 1 | The cortical gray mantle is thinned |
| Dentate hilus discoloration | 0 | The cerebellar sections show normal appearance of the dentate hilus |
|  | 1 | The cerebellar sections show gray discoloration of the dentate hilus |
| Dentate nucleus atrophy | 0 | The dentate nucleus is unremarkable |
|  | 1 | The dentate nucleus shows atrophy |
| Dorsal leptomeninges abnormality | 0 | The dorsal leptomeninges are clear |
|  | 1 | The dorsal leptomeninges have focal fibrous thickening |
| Fornix atrophy | 0 | The fornix is unremarkable |
|  | 1 | The fornix is thin and discolored |
| Fourth ventricle enlargement | 0 | The infratentorial structures show no obvious enlargement of the fourth ventricle |
|  | 1 | The infratentorial structures show enlargement of the fourth ventricle |
|  | 2 | The infratentorial structures show marked enlargement of the fourth ventricle |
| Frontal convexity atrophy | 0 | The sulci and gyri reveal no atrophy over the frontal convexity |
|  | 1 | The sulci and gyri reveal atrophy over the frontal convexity |
|  | 2 | The sulci and gyri reveal marked atrophy over the frontal convexity |
| Frontal horn enlargement | 0 | Sequential sections reveal no enlargement of the frontal horn of the lateral ventricle |
|  | 1 | Sequential sections reveal enlargement of the frontal horn of the lateral ventricle |
| Frontal lobe atrophy | 0 | The sulci and gyri reveal no cortical atrophy over the frontal lobe |
|  | 1 | The sulci and gyri reveal cortical atrophy over the frontal lobe |
|  | 2 | The sulci and gyri reveal marked cortical atrophy over the frontal lobe |
| Globus pallidus atrophy | 0 | Basal ganglia show no atrophy of the globus pallidus |
|  | 1 | The basal ganglia show atrophy of the globus pallidus |
|  | 2 | The basal ganglia show marked atrophy of the globus pallidus |
| Globus pallidus discoloration | 0 | The basal ganglia show no discoloration of the globus pallidus |
|  | 1 | The basal ganglia show discoloration of the globus pallidus |
| Hippocampal atrophy | 0 | The hippocampal formation is unremarkable |
|  | 1 | The hippocampal formation is atrophic |
|  | 2 | The hippocampal formation has marked atrophy |
| Locus coeruleus depigmentation | 0 | There is visible pigment in the locus ceruleus |
|  | 1 | The locus ceruleus has decreased pigmentation |
| Medial temporal lobe atrophy | 0 | The medial temporal lobe has no atrophy |
|  | 1 | The medial temporal lobe has atrophy |
|  | 2 | The medial temporal lobe is markedly atrophic |
| Midbrain atrophy | 0 | Horizontal sections of the midbrain at right angles to the neuraxis are unremarkable |
|  | 1 | Horizontal sections of the midbrain at right angles to the neuraxis are remarkable for midbrain atrophy |
| Occipital lobe atrophy | 0 | There is no significant atrophy of occipital lobe |
|  | 1 | The sulci and gyri reveal cortical atrophy over the occipital lobe |
|  | 2 | The sulci and gyri reveal marked cortical atrophy over the occipital lobe |
| Parietal lobe atrophy | 0 | There is no significant atrophy of parietal lobe |
|  | 1 | The sulci and gyri reveal cortical atrophy over the parietal lobe |
|  | 2 | The sulci and gyri reveal marked cortical atrophy over the parietal lobe |
| Pontine atrophy | 0 | Horizontal sections of the pons at right angles to the neuraxis are unremarkable |
|  | 1 | Horizontal sections of the pons at right angles to the neuraxis are remarkable for pontine atrophy |
| Posterior cortical atrophy | 0 | There is no atrophy of the posterior cortical areas |
|  | 1 | The sulci and gyri reveal atrophy over the posterior cortical areas |
|  | 2 | The sulci and gyri reveal marked atrophy over the posterior cortical areas |
| Putamen atrophy | 0 | The basal ganglia show no atrophy of the putamen |
|  | 1 | The basal ganglia show atrophy of the putamen |
|  | 2 | The basal ganglia show marked atrophy of the putamen |
| Putamen discoloration | 0 | The basal ganglia show no discoloration of the putamen |
|  | 1 | The basal ganglia show discoloration of the putamen |
| Striatum atrophy | 0 | The striatum is unremarkable |
|  | 1 | The striatum is atrophic |
|  | 2 | The striatum is markedly atrophic |
| Substantia nigra depigmentation | 0 | The substantia nigra has visible pigmentation |
|  | 1 | The substantia nigra has decreased pigmentation |
|  | 2 | The substantia nigra has markedly decreased pigmentation |
| Subthalamic nucleus atrophy | 0 | There is no atrophy of the subthalamic nucleus |
|  | 1 | The subthalamic nucleus has atrophy |
|  | 2 | The subthalamic nucleus has marked atrophy |
| Superior cerebellar peduncle atrophy | 0 | The superior cerebellar peduncle is preserved |
|  | 1 | The superior cerebellar peduncle has atrophy |
|  | 2 | The superior cerebellar peduncle has marked atrophy |
| Temporal horn enlargement | 0 | Sequential sections reveal no enlargement of the temporal horn of the lateral ventricle |
|  | 1 | Sequential sections reveal enlargement of the temporal horn of the lateral ventricle |
| Temporal lobe atrophy | 0 | The temporal lobe has no atrophy |
|  | 1 | The sulci and gyri reveal cortical atrophy over the temporal lobe |
|  | 2 | The sulci and gyri reveal marked cortical atrophy over the temporal lobe |
| Thalamic atrophy | 0 | The thalamus is unremarkable |
|  | 1 | The thalamus is atrophic |
| Ventricular system abnormality | 0 | The ventricular system is undilated |
|  | 1 | The ventricular system is dilated |

**Supplementary Table S2** Prompts Used for Fine-Tuning and Prediction with an LLM

1. Prompt for Scoring Macroscopic Findings

You are an expert neuropathologist. Based on the gross description below, classify the presence/severity of {finding} on a scale of {range}. If it cannot be determined, answer 'NA'.

2. Prompt for Text-Based Classification

Make a diagnosis on the basis of the gross findings provided below.

{Age}, {Sex}, {Brain_weight}, {macrofindings}

{finding} and {range} correspond to the “Findings” and “Score” columns, respectively, **in Supplementary Table S1.**

**Supplementary Table S3** Macroscopic Findings and Feature Importance by Neuropathologic Diagnosis

|  | Overall | AD | AD-LBD | LBD | FTLD | CBD | PSP | MSA | P value | Feature importance |
| --- | --- | --- | --- | --- | --- | --- | --- | --- | --- | --- |
|  | 5613 | 1677 | 746 | 856 | 270 | 280 | 1450 | 334 |  |  |
| Male, n (%) | 3075 (54.8) | 771 (46.0) | 412 (55.2) | 625 (73.0) | 150 (55.6) | 144 (51.4) | 805 (55.5) | 168 (50.3) | **<0.01** | 2.86 |
| Age at death (y) | 76 [69, 82] | 80 [72, 86] | 78 [73, 84] | 77 [72, 82] | 70 [63, 77] | 69 [65, 74] | 74 [69, 80] | 66 [60, 71] | **<0.01** | 10.40 |
| Brain weight (g) | 1100 [1000, 1220] | 1020 [920, 1120] | 1100 [980, 1200] | 1200 [1100, 1300] | 1000 [900, 1120] | 1100 [1000, 1200] | 1140 [1040, 1240] | 1200 [1100, 1300] | **<0.01** | 9.31 |
| Cortical atrophy | 3559 (63.4%) | 1484 (88.5%) | 649 (87.0%) | 583 (68.1%) | 226 (83.7%) | 195 (69.6%) | 378 (26.1%) | 44 (13.2%) | **<0.01** | 4.27 |
| Frontal convexity atrophy | 1527 (27.2%) | 319 (19.0%) | 160 (21.4%) | 240 (28.0%) | 45 (16.7%) | 55 (19.6%) | 696 (48.0%) | 12 (3.6%) | **<0.01** | 2.32 |
| Anterior cortical atrophy | 211 (3.8%) | 93 (5.5%) | 37 (5.0%) | 10 (1.2%) | 47 (17.4%) | 5 (1.8%) | 15 (1.0%) | 4 (1.2%) | **<0.01** | 0.45 |
| Posterior cortical atrophy | 363 (6.5%) | 156 (9.3%) | 51 (6.8%) | 19 (2.2%) | 17 (6.3%) | 16 (5.7%) | 102 (7.0%) | 2 (0.6%) | **<0.01** | 0.43 |
| Frontal lobe atrophy | 1382 (24.6%) | 571 (34.0%) | 204 (27.3%) | 94 (11.0%) | 173 (64.1%) | 61 (21.8%) | 260 (17.9%) | 19 (5.7%) | **<0.01** | 2.42 |
| Parietal lobe atrophy | 727 (13.0%) | 440 (26.2%) | 131 (17.6%) | 30 (3.5%) | 86 (31.9%) | 22 (7.9%) | 17 (1.2%) | 1 (0.3%) | **<0.01** | 0.88 |
| Temporal lobe atrophy | 2024 (36.1%) | 1276 (76.1%) | 420 (56.3%) | 83 (9.7%) | 163 (60.4%) | 28 (10.0%) | 49 (3.4%) | 5 (1.5%) | **<0.01** | 2.41 |
| Medial temporal lobe atrophy | 1909 (34.0%) | 1239 (73.9%) | 403 (54.0%) | 69 (8.1%) | 141 (52.2%) | 23 (8.2%) | 33 (2.3%) | 1 (0.3%) | **<0.01** | 3.65 |
| Occipital lobe atrophy | 654 (11.7%) | 420 (25.0%) | 119 (16.0%) | 26 (3.0%) | 57 (21.1%) | 19 (6.8%) | 9 (0.6%) | 4 (1.2%) | **<0.01** | 0.65 |
| Cortical gray mantle thinning | 1524 (27.2%) | 803 (47.9%) | 278 (37.3%) | 27 (3.2%) | 184 (68.1%) | 146 (52.1%) | 84 (5.8%) | 2 (0.6%) | **<0.01** | 3.73 |
| Cerebral white matter abnormality | 2064 (36.8%) | 1004 (59.9%) | 326 (43.7%) | 156 (18.2%) | 159 (58.9%) | 166 (59.3%) | 225 (15.5%) | 28 (8.4%) | **<0.01** | 3.55 |
| Corpus callosum thinning | 273 (4.9%) | 34 (2.0%) | 12 (1.6%) | 4 (0.5%) | 31 (11.5%) | 105 (37.5%) | 86 (5.9%) | 1 (0.3%) | **<0.01** | 2.36 |
| Ventricular system abnormality | 1512 (26.9%) | 320 (19.1%) | 149 (20.0%) | 190 (22.2%) | 46 (17.0%) | 66 (23.6%) | 513 (35.4%) | 228 (68.3%) | **<0.01** | 1.95 |
| Frontal horn enlargement | 4044 (72.0%) | 1130 (67.4%) | 516 (69.2%) | 533 (62.3%) | 218 (80.7%) | 233 (83.2%) | 1141 (78.7%) | 273 (81.7%) | **<0.01** | 2.82 |
| Temporal horn enlargement | 2008 (35.8%) | 966 (57.6%) | 424 (56.8%) | 279 (32.6%) | 158 (58.5%) | 40 (14.3%) | 133 (9.2%) | 8 (2.4%) | **<0.01** | 3.78 |
| Fourth ventricle enlargement | 1060 (18.9%) | 111 (6.6%) | 39 (5.2%) | 47 (5.5%) | 21 (7.8%) | 49 (17.5%) | 611 (42.1%) | 182 (54.5%) | **<0.01** | 0.71 |
| Hippocampal atrophy | 2257 (40.2%) | 1255 (74.8%) | 505 (67.7%) | 249 (29.1%) | 170 (63.0%) | 27 (9.6%) | 44 (3.0%) | 7 (2.1%) | **<0.01** | 3.39 |
| Amygdala atrophy | 2247 (40.0%) | 1256 (74.9%) | 499 (66.9%) | 245 (28.6%) | 173 (64.1%) | 27 (9.6%) | 41 (2.8%) | 6 (1.8%) | **<0.01** | 2.82 |
| Fornix atrophy | 23 (0.4%) | 11 (0.7%) | 5 (0.7%) | 0 (0.0%) | 7 (2.6%) | 0 (0.0%) | 0 (0.0%) | 0 (0.0%) | **<0.01** | 0.06 |
| Anterior commissure abnormality | 325 (5.8%) | 5 (0.3%) | 4 (0.5%) | 7 (0.8%) | 8 (3.0%) | 28 (10.0%) | 270 (18.6%) | 3 (0.9%) | **<0.01** | 0.11 |
| Basal ganglia atrophy | 844 (15.0%) | 78 (4.7%) | 38 (5.1%) | 19 (2.2%) | 121 (44.8%) | 135 (48.2%) | 154 (10.6%) | 299 (89.5%) | **<0.01** | 1.75 |
| Striatum atrophy | 3 (0.1%) | 0 (0.0%) | 0 (0.0%) | 0 (0.0%) | 1 (0.4%) | 0 (0.0%) | 1 (0.1%) | 1 (0.3%) | 0.09 | 0.00 |
| Caudate atrophy | 359 (6.4%) | 74 (4.4%) | 34 (4.6%) | 8 (0.9%) | 112 (41.5%) | 100 (35.7%) | 30 (2.1%) | 1 (0.3%) | **<0.01** | 1.97 |
| Putamen atrophy | 377 (6.7%) | 11 (0.7%) | 4 (0.5%) | 12 (1.4%) | 2 (0.7%) | 31 (11.1%) | 18 (1.2%) | 299 (89.5%) | **<0.01** | 3.42 |
| Putamen discoloration | 377 (6.7%) | 11 (0.7%) | 4 (0.5%) | 12 (1.4%) | 2 (0.7%) | 31 (11.1%) | 18 (1.2%) | 299 (89.5%) | **<0.01** | 2.16 |
| Globus pallidus atrophy | 325 (5.8%) | 10 (0.6%) | 7 (0.9%) | 11 (1.3%) | 37 (13.7%) | 118 (42.1%) | 133 (9.2%) | 9 (2.7%) | **<0.01** | 0.51 |
| Globus pallidus discoloration | 325 (5.8%) | 10 (0.6%) | 7 (0.9%) | 11 (1.3%) | 37 (13.7%) | 118 (42.1%) | 133 (9.2%) | 9 (2.7%) | **<0.01** | 0.41 |
| Thalamic atrophy | 212 (3.8%) | 48 (2.9%) | 25 (3.4%) | 3 (0.4%) | 66 (24.4%) | 40 (14.3%) | 29 (2.0%) | 1 (0.3%) | **<0.01** | 0.41 |
| Subthalamic nucleus atrophy | 1417 (25.2%) | 9 (0.5%) | 12 (1.6%) | 24 (2.8%) | 19 (7.0%) | 87 (31.1%) | 1263 (87.1%) | 3 (0.9%) | **<0.01** | 2.64 |
| Midbrain atrophy | 1700 (30.3%) | 35 (2.1%) | 25 (3.4%) | 34 (4.0%) | 81 (30.0%) | 101 (36.1%) | 1179 (81.3%) | 245 (73.4%) | **<0.01** | 1.93 |
| Pontine atrophy | 1638 (29.2%) | 28 (1.7%) | 23 (3.1%) | 23 (2.7%) | 73 (27.0%) | 84 (30.0%) | 1179 (81.3%) | 228 (68.3%) | **<0.01** | 1.30 |
| Substantia nigra depigmentation | 3777 (67.3%) | 548 (32.7%) | 500 (67.0%) | 758 (88.6%) | 138 (51.1%) | 222 (79.3%) | 1308 (90.2%) | 303 (90.7%) | **<0.01** | 4.51 |
| Locus coeruleus depigmentation | 3488 (62.1%) | 550 (32.8%) | 489 (65.5%) | 743 (86.8%) | 103 (38.1%) | 183 (65.4%) | 1217 (83.9%) | 203 (60.8%) | **<0.01** | 4.23 |
| Superior cerebellar peduncle atrophy | 1302 (23.2%) | 4 (0.2%) | 7 (0.9%) | 15 (1.8%) | 4 (1.5%) | 66 (23.6%) | 1203 (83.0%) | 3 (0.9%) | **<0.01** | 1.72 |
| Dentate nucleus atrophy | 1298 (23.1%) | 13 (0.8%) | 13 (1.7%) | 17 (2.0%) | 5 (1.9%) | 56 (20.0%) | 1186 (81.8%) | 8 (2.4%) | **<0.01** | 2.20 |
| Dentate hilus discoloration | 1293 (23.0%) | 18 (1.1%) | 15 (2.0%) | 20 (2.3%) | 7 (2.6%) | 62 (22.1%) | 1168 (80.6%) | 3 (0.9%) | **<0.01** | 0.91 |
| Cerebellar white matter atrophy | 87 (1.5%) | 1 (0.1%) | 2 (0.3%) | 3 (0.4%) | 0 (0.0%) | 3 (1.1%) | 11 (0.8%) | 67 (20.1%) | **<0.01** | 0.13 |
| Atherosclerosis | 2180 (38.8%) | 988 (58.9%) | 370 (49.6%) | 354 (41.4%) | 96 (35.6%) | 60 (21.4%) | 279 (19.2%) | 33 (9.9%) | **<0.01** | 3.97 |
| Dorsal leptomeninges abnormality | 404 (7.2%) | 170 (10.1%) | 53 (7.1%) | 39 (4.6%) | 16 (5.9%) | 13 (4.6%) | 105 (7.2%) | 8 (2.4%) | **<0.01** | 0.50 |

Values are median [interquartile range] or n (%). Percentages for gross findings use the full diagnostic group as the denominator. Gross findings were classified as absent (score 0) or present (score ≥1), irrespective of severity. Feature importance values refer to the CatBoost score-based model and sum to 100 across the 42 model inputs. Abbreviations: AD, Alzheimer disease; AD-LBD, combined Alzheimer disease and Lewy body disease; CBD, corticobasal degeneration; FTLD, frontotemporal lobar degeneration; LBD, Lewy body disease; MSA, multiple system atrophy; PSP, progressive supranuclear palsy.
